# Caffeine and Methylliberine: A Human Pharmacokinetic Interaction Study

**DOI:** 10.1101/2021.01.05.21249234

**Authors:** Goutam Mondal, Yan-Hong Wang, ZaChara Catchings, Matthew Butawan, Richard J. Bloomer, Charles R. Yates

**Affiliations:** The National Center for Natural Products Research, University of Mississippi, Oxford, MS, USA; Cardiorespiratory/Metabolic Laboratory, School of Health Studies, University of Memphis, Memphis, TN, USA; NPI, LLC, Oxford, MS

**Keywords:** caffeine, methylliberine, theacrine, pharmacokinetics

## Abstract

Methylliberine and theacrine are methylurates found in the leaves of various *Coffea* species and *Camellia assamica* var. *kucha*, respectively. We previously demonstrated that the methylxanthine caffeine increased theacrine’s oral bioavailability in humans. Consequently, we conducted a double-blind, placebo-controlled study pharmacokinetic study in humans administered methylliberine, theacrine, and caffeine to determine methylliberine’s pharmacokinetic interaction potential with either caffeine or theacrine. Subjects (n = 12) received an oral dose of either methylliberine (25 or 100 mg), caffeine (150 mg), methylliberine (100 mg) plus caffeine (150 mg), or methylliberine (100 mg) plus theacrine (50 mg) using a randomized, double-blind, crossover design. Blood samples were collected over 24 hours and analyzed for methylliberine, theacrine, and caffeine using UPLC-MS/MS. Methylliberine exhibited linear pharmacokinetics that were unaffected by co-administration of either caffeine or theacrine. However, methylliberine co-administration resulted in decreased oral clearance (41.9 ± 19.5 vs. 17.1 ± 7.80 L/hr) and increased half-life (7.2 ± 5.6 versus 15 ± 5.8 hrs) of caffeine. Methylliberine had no impact on caffeine’s maximum concentration (440 ± 140 vs. 458 ± 93.5 ng/mL) or oral volume of distribution (351 ± 148 vs. 316 ± 76.4 L). We previously demonstrated theacrine bioavailability was enhanced by caffeine, however, caffeine pharmacokinetics were unaffected by theacrine. Herein, we found that methylliberine altered caffeine pharmacokinetics without a reciprocal interaction, which suggests caffeine may interact uniquely with different methylurates. Understanding the mechanism(s) of interaction between methylxanthines and methylurates is of critical importance in light of the recent advent of dietary supplements containing both purine alkaloid classes.

## Introduction

The methylxanthine caffeine is found across the globe in a wide variety of plant genera including *Camellia* (e.g., *C. sinensis*), *Coffea* (e.g., *C. arabica*), *Cola* (e.g., *C. nitida*), *Paullinia* (e.g., *P. cupana*), and *Ilex* (e.g., *I. paraguarensis*)^1^. Plants that produce purine alkaloids such as caffeine have been prized for thousands of years for their medicinal properties. Today, caffeine is potentially the most highly consumed phytochemical as a consequence of its pervasive utilization in foods, beverages, and dietary supplements. A prime example being caffeine-containing energy drinks and shots, which are popular among all age groups due to their ability to enhance energy, mood, and focus (EMF) ^2,3^. When consumed responsibly, caffeine is associated with relatively few adverse events that include tolerance, disrupted sleep, and withdrawal. However, the limitations of caffeine use by some has led to the exploration for unique purine alkaloids as caffeine alternatives to enhance EMF. Indeed, phytochemical characterization of unique *Coffea* ^4^ and *Camellia* species ^5^ has greatly expanded the field of research into pharmacologic alternatives to caffeine.

Young leaves, pericarp, and seeds of *C. liberica* were found to contain the methylurate theacrine (1,3,7,9-tetramethyluric acid)^4^, which confirmed the first description of theacrine in a tea plant ^6^. Later, this same research group identified theacrine in cupu seeds (Theobroma grandiflorum) ^7^. More recently, the kudingcha tea plant (*C. assamica* var. *kucha*), found in the Chinese province of Yunnan, was shown to uniquely produce theacrine (**Figure 1**) as it primary purine alkaloid ^5^. Radioactive caffeine tracer studies designed to explore purine metabolism in the leaves of various *Coffea* species demonstrated that during stage 1 of vegetative development, young plants accumulated caffeine synthesized from theobromine ^8^. In stage 2, caffeine is gradually converted to theacrine, which is then converted in stage 3 to liberine (O(2),1, 9-tetramethyluric acid) (stage 3), presumably through the intermediate metabolite methylliberine (O(2),1,7,9-tetramethyluric acid). In *C. assamica* var. *kucha* leaves, pulse-chase experiments using [8-^14^C]adenosine demonstrated that theacrine was synthesized from caffeine in a purported three-step pathway involving the intermediate 1,3,7-methyluric acid. However, neither methylliberine nor liberine was detected ^5^.

**Figure 1.**
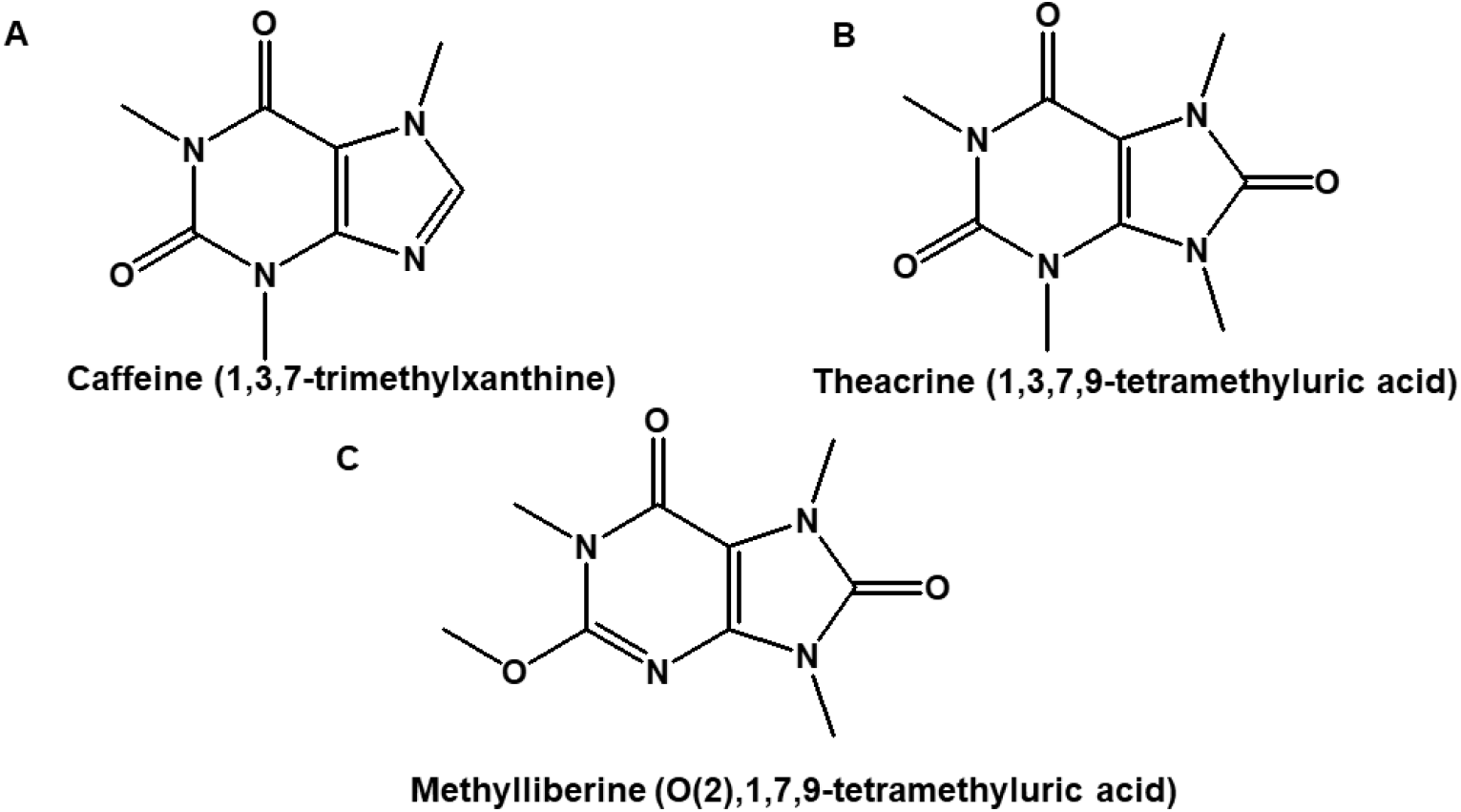
Chemical structures of (A) caffeine, (B) theacrine, and (C) methylliberine.

Studies have shown that theacrine, similar to caffeine, exerts psychostimulatory action via modulation of the adenosinergic and dopaminergic pathways ^9^. Unlike caffeine, however, theacrine does not appear to be associated with tolerance, nor does it negatively affect the cardiovascular system ^9^. Consequently, theacrine has been evaluated as a means to augment, and thus potentially reduce the dose of, caffeine in clinical substantiation studies for EMF. In humans, co-administration of theacrine (125 mg) and caffeine (150 mg), compared to caffeine alone (275 mg) or placebo, resulted in sustained focus and concentration under fatigue-inducing conditions as well as enhanced alertness, attention, and information processing ^3^. To elucidate the potential mechanism(s) underlying theacrine’s hypothesized pharmacologic augmentation, we conducted a randomized, double-blind crossover study in humans to determine whether a pharmacokinetic interaction existed between theacrine and caffeine ^9^. Interestingly, we found that theacrine oral bioavailability was enhanced by caffeine, however, caffeine pharmacokinetics were unaffected by theacrine. Thus, contrary to our original hypothesis, it seems more likely that caffeine enhanced theacrine pharmacodynamics secondary to increased theacrine plasma exposure.

Following on the heels of theacrine’s commercial success in the dietary supplement arena, methylliberine has recently been granted new dietary ingredient (NDI) status following completion of a 90-day repeated-dose oral toxicity study as required by the Dietary Supplement Health Education Act ^10^. In addition, human adverse event potential studies using methylliberine alone, and in combination with theacrine, found no effect of methylliberine on heart rhythm (electrocardiogram; ECG), resting heart rate, or blood pressure ^11^. Additionally, anecdotal benefits of methylliberine suggesting reduced onset of action of EMF activity without anxiety has led to methlliberine being “stacked” (i.e., combined) with caffeine and/or theacrine. Because we previously demonstrated the interaction potential between the methylxanthine caffeine and the methylurate theacrine, we hypothesized that caffeine would interact with the methylurate methylliberine. Therefore, the purpose of this study was to determine methylliberine pharmacokinetics and its pharmacokinetic interaction potential with theacrine and caffeine following oral administration to humans.

## Methods

The study protocol and informed consent were approved by the University of Memphis Institutional Review Board. Study participants were informed of all procedures, potential benefits, and risks associated with the study and provided informed consent prior to any study-related procedures. The clinical study was conducted at the University of Memphis in accordance with the US Code of Federal Regulations (CFR) governing Protection of Human Subjects (21 CFR Part 50), Financial Disclosure by Clinical Investigator (21 CFR Part 54), and Institutional Review Board (IRB) (21 CFR Part 56). Moreover, the study adhered to the 1996 guidelines of the International Conference on Harmonization (Good Clinical Practice (GCP)), which is consistent with the Declaration of Helsinki as adopted in 2008.

### Study Design

Study description and eligbility were previously described^12^. In brief, this was a randomized, double-blind, crossover study designed to assess the pharmacokinetic interaction potential of methylliberine, caffeine, and theacrine in healthy subjects. Subjects were randomized using a 4×4 Latin Square design to receive a single oral dose of either methylliberine 25 mg (low dose), methylliberine 100 mg (high dose), caffeine 150 mg, methylliberine 100 mg and caffeine 150 mg or methylliberine 100 mg and theacrine 50 mg. Methylliberine (Dynamine®) and theacrine (TeaCrine®) were provided by Compound Solutions (Carlsbad, CA). Caffeine, administered as caffeine anhydrous, was obtained from Nutravative Ingredients (Allen, TX).

### LC-MS/MS

Plasma levels of caffeine, methylliberine, and theacrine were measured using a previously described UPLC-MS/MS method^13^. Briefly, bioanalytical method inter- and intra-day accuracy and precision were verified to be ±15%. The lower limit of quantification for caffeine, methylliberine, and theacrine was 0.67 ng/mL. Plasma samples were extracted with methanol containing the internal standard (caffeine-^13^C_3_). The LC-MS/MS system comprised a Waters Acquity UPLC™ I-class system (Waters Corporation, Milford, MA, USA) coupled with a Xevo TQ-S triple quadrupole mass spectrometry detector operating in the negative electrospray ionization (ESI) mode (capillary voltage, 1.1 kV; source temperature, 150 °C; desolvation temperature, 500 °C; desolvation gas flow, 1000 L/h, and cone gas flow, 150 L/h). Separation was achieved using an UPLC BEH C_18_ column (50 mm × 2.1 mm I.D., 1.8 µm) and a mobile phase comprising water containing 0.1 % formic acid (A) and acetonitrile with 0.1 % formic acid (B). Detection was obtained using the Multiple Reaction Monitoring (MRM) mode including two MRMs for confirmation of each analyte. The quantification MRMs for caffeine, caffeine-^13^C_3_ (IS), theacrine, and methylliberine were m/z 195.11→138.01, 198.1→140.07, 225.12→168.02, and 225.12→167.95, respectively.

### Pharmacokinetic data analysis

Caffeine, methylliberine, and theacrine oral pharmacokinetic parameters were estimated from plasma concentration-time data, adjusted for lag time (t_lag_), using noncompartmental methods in Phoenix WinNonlin (version 8.2, Certara USA, Inc., Princeton, NJ). Maximum concentration (C_max_) and time corresponding to C_max_ (T_max_) were determined from the plasma concentration versus time data. Area under the plasma concentration-time curve from time 0 to infinity (AUC_0–∞_) was calculated using the linear trapezoidal rule. The terminal half-life (t_1/2_), was evaluated using ln 2/k_el_, with k_el_ as the terminal rate elimination constant estimated from the slope of the linear regression of the log plasma concentration versus time curve during the terminal phase. The oral clearance (CL/F) was calculated by dividing the administered oral dose by AUC0-∞. The apparent oral volume of distribution during the terminal elimination phase (Vz/F) was calculated by dividing CL/F by k_el_.

### Statistical analysis

To determine the probability of interaction magnitude between methylliberine and caffeine and/or theacrine, pharmacokinetic parameters were first logarithmically transformed. For each parameter, mean differences of the transformed values were obtained by taking the average of the difference of the transformed values, and upper and lower confident level with a 90% confidence interval (CI) were obtained using the paired *t* test function in Excel. The results of this analysis were exponentiated which corresponded to 90% confidence intervals around the geometric mean ratios for any observed pharmacokinetic parameters^14^.

## Results

### Subject characteristics

Twelve healthy men (n=6; aged -- years; -------kg) and women (n=6; aged------years; -----kg) participated in this study. Men and women ingested a daily amount of caffeine mg and ---mg respectively. All subjects were well tolerated all treatments and no adverse effect was recorded. Diet intake was not changed across all treatment conditions for total kilocalorie, macro-and micro-nutrient composition.

### Pharmacokinetics

Mean plasma concentration (± standard deviation) time profiles for methylliberine, caffeine, and theacrine are shown in **Figures 2A-C, 3, 4 and Figure S1**. Methylliberine pharmacokinetic parameters for each cohort are shown in **Table 1**. We found the methylliberine was rapidly absorbed from the oral administration, with C_max_ reached on average at 0.6 and 0.9 hours after following low (25 mg) and high (100 mg) doses of methylliberine, respectively (**Table 1, Figure 2A**). Thereafter, methylliberine was eliminated with a half-life averaging 1 to 1.4 hours. Dose-normalized C_max_ and AUC were significantly higher, oral clearance and oral volume of distribution were significantly lower, following 100 mg dose of methylliberine compared to 25 mg of methylliberine.

**Figure 2.**
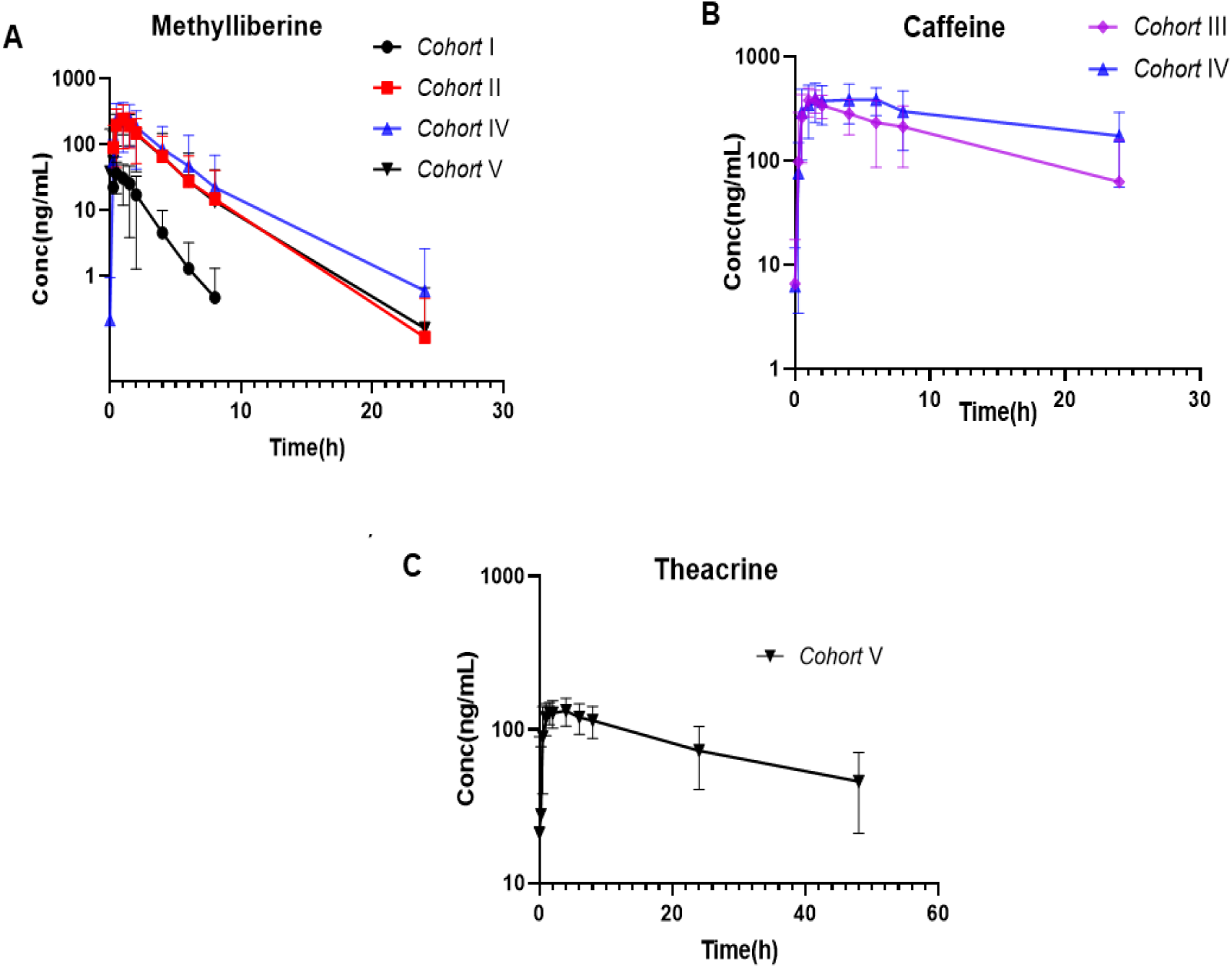
Plasma concentrations-time profile of (A) methylliberine in Cohort I, cohort II, cohort IV, and cohort V; (B) caffeine in cohort III, and cohort IV; and (C) theacrine in cohort V. Data represented as the mean ± SD. Cohort I, methylliberine 25 mg; cohort II, methylliberine 100 mg; cohort III, caffeine 150 mg; cohort IV, methylliberine 100 mg and caffeine 150 mg; cohort V, methylliberine 100 mg and theacrine 50 mg; SD, standard deviation.

**Figure 3.**
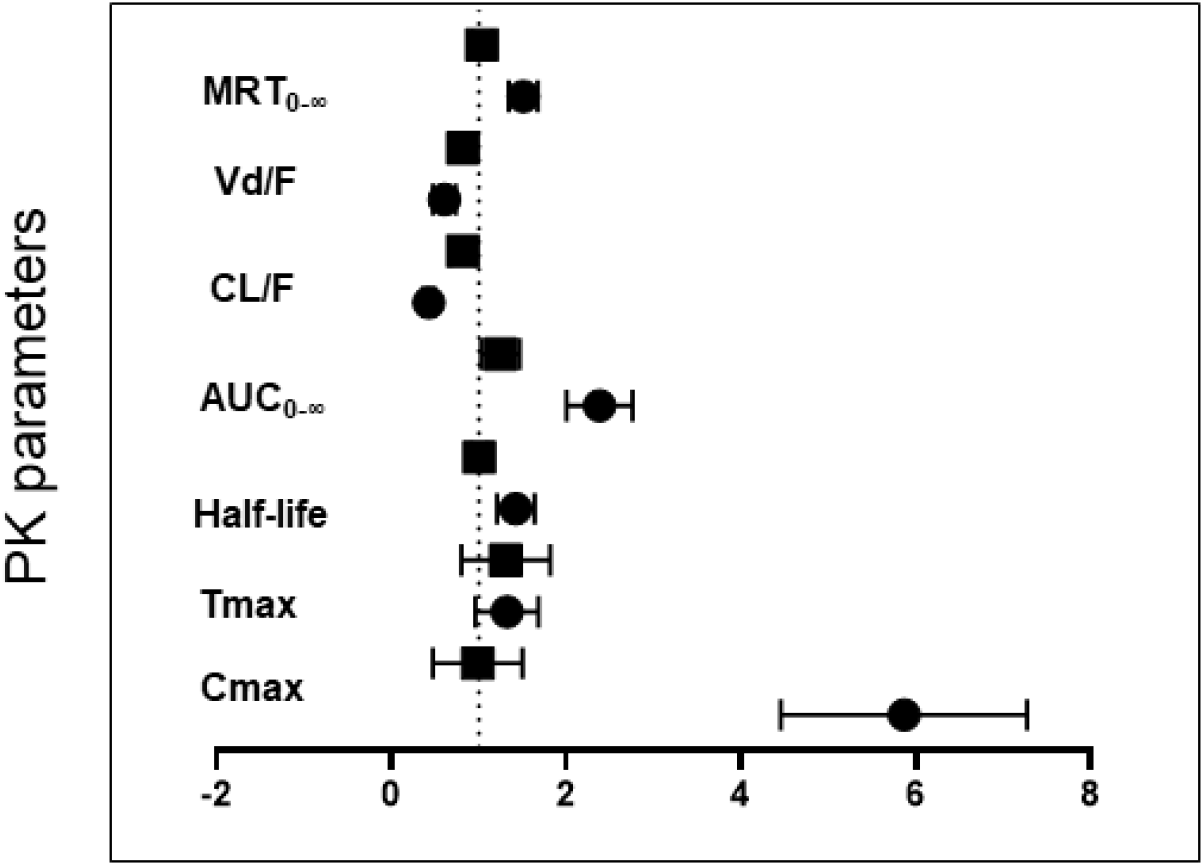
Forest plot demonstrating the degree of probability of interaction between methylliberine with caffeine and/or theacrine using 90% confidence intervals about the geometric mean ratio of the observed pharmacokinetic parameters following a single oral methylliberine dose (•25 mg, ▪ 100 mg) alone or in combination with caffeine (150 mg). MRT_0-∞_, mean residence time zero to infinity; CL/F, oral clearance; Vd/F, oral volume of distribution; AUC_0-∞_, area under the curve from zero to time infinity (dose normalized); Cmax, maximum plasma concentration (dose normalized); Tmax, time to reach maximum plasma concentration.

**Figure 4.**
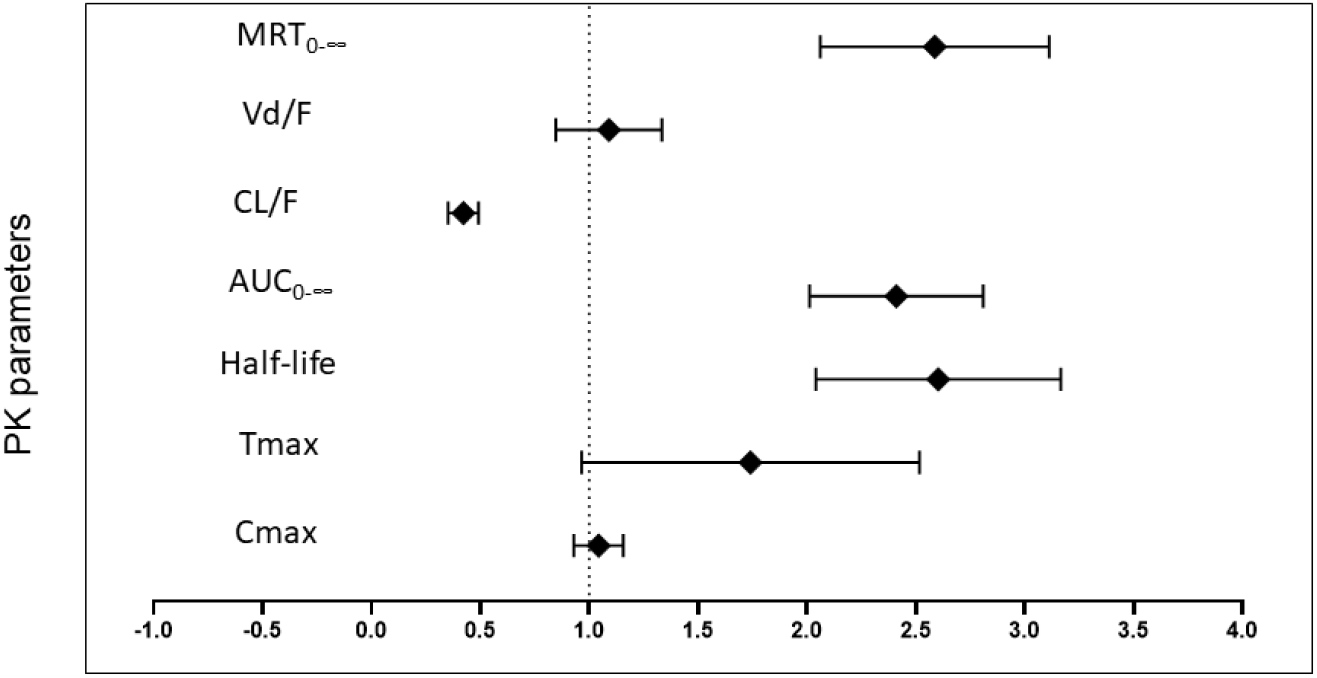
Forest plot demonstrating the degree of probability of interaction between methylliberine and caffeine using 90% confidence intervals about the geometric mean ratio of the observed pharmacokinetic parameters following a single oral caffeine dose (150 mg) alone or in combination with methylliberine (100 mg). MRT_0-∞_, mean residence time zero to infinity; CL/F, oral clearance; Vd/F, oral volume of distribution; AUC_0-∞_, area under the curve from zero to time infinity (dose normalized); Cmax, maximum plasma concentration (dose normalized); Tmax, time to reach maximum plasma concentration.

**TABLE 1.** Methylliberine pharmacokinetic parameters

| *Parameter* | *Cohort I* | *Cohort II* | *Cohort IV* | *Cohort V* |
| --- | --- | --- | --- | --- |
| $C_{\max}$ (ng/mL) | 55.3±34.6 | 287±141 | 349±130 | 288.6±91.6 |
| $T_{\max}$ (hours) | 0.6±0.3 | 0.9±0.3 | 0.9±0.4 | 0.8±0.5 |
| $t_{1/2}$ (hours) | 1.0±0.3 | 1.4±0.6 | 1.5±0.8 | 1.4±0.6 |
| AUC (h x ng/<br>mL/mg) | 3.4±2.2 | 8.0±6.8 | 11.2±11.6 | 7.6±6.6 |
| CL/F (L/h) | 425.8±261.5 | 201.2±121.7 | 148.7±88.9 | 189.3±87.6 |
| $V_d/F$ (L) | 556.2±253.7 | 356.4±163.9 | 270.2±126.3 | 316.4±99.6 |
| MRT (hours) | 1.6±0.5 | 2.4±1 | 2.6±1.2 | 2.3±1.1 |

After a single dose of methylliberine 25 mg, the geometric mean Cmax was 44.16 ng/mL, t_1/2_ was 0.99 h, AUC was 2.83 ng·h/mL/mg, CL/F was 352.9 L/h, V_d_/F was 505.6 L (**Table S1**). Compared with methylliberine 25 mg, oral administration of methylliberine 100 mg resulted in an increase in the geometric mean C_max_ was 254.09 ng/mL, AUC was 6.69 ng·h/mL/mg, MRT was 2.31 h, and in a decrease in the geometric mean V_d_/F was 302.6 L, CL/F was 149.4 L/h (**Table S1**). The geometric mean ratios for C_max_, T_max_, half-life, AUC, CL/F, Vd/F, and MRT on oral administration of methylliberine (100 mg) versus methylliberine (25 mg) were 5.75, 1.29, 1.41, 2.36, 0.42, 0.6, and 1.5 respectively **Table S1**. Based on the geometric means of Cmax, Tmax,, AUC, CL/F, and Vd/F, exposure of methylliberine (100 mg) was different than methylliberine and when coadministered with caffeine **Table 2**, but was comparable between methylliberine and when coadministered with theacrine **Table S2**. The geometric ratios for C_max_, T_max_, half-life, AUC, CL/F, Vd/F, and MRT on oral coadministration of methylliberine (100 mg) with caffeine (150 mg) versus methlliberine (100 mg) alone were 0.9, 1.24, 1.0, 1.23, 0.81, 0.81, and 1.03 respectively **Table 2**. The geometric ratios for C_max_, T_max_, half-life, AUC, CL/F, Vd/F, and MRT on oral coadministration of methylliberine (100 mg) with theacrine (50 mg) versus methlliberine (100 mg) alone were 1.08, 1.03, 0.95, 0.95, 1.05, 0.99, and 1.03 respectively **Table S2**.

**TABLE 2.**
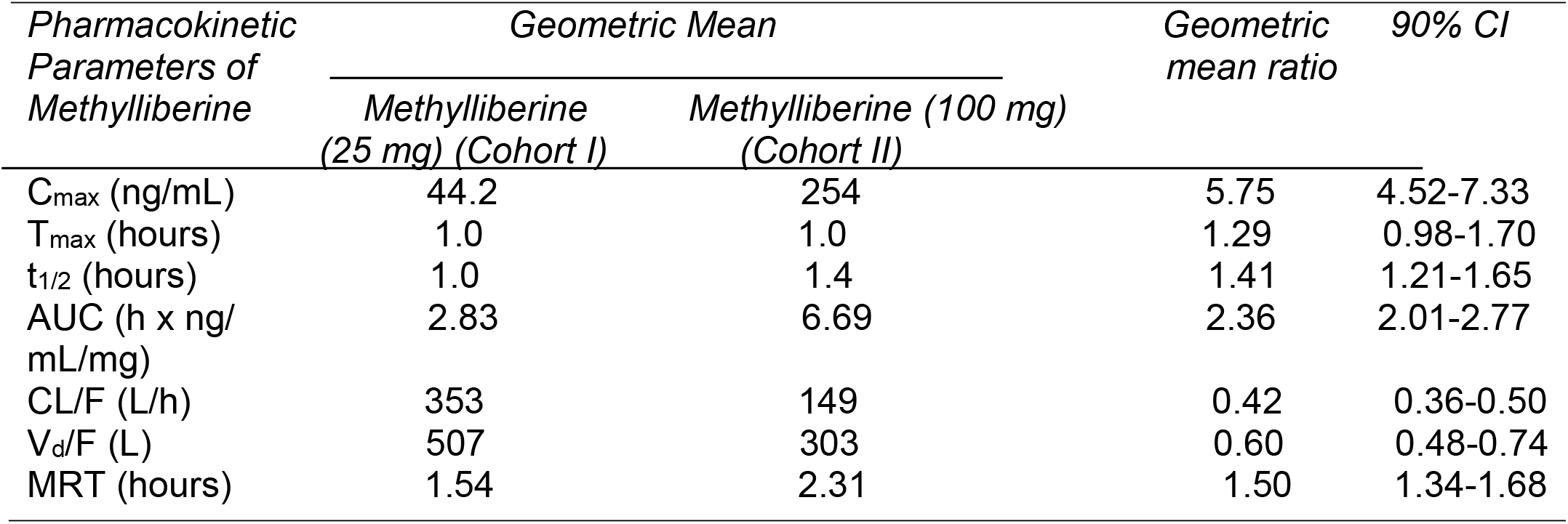
Summary of the statistical analysis of the pharmacokinetic parameters of methylliberine after oral administration of a 25 mg dose and 100 mg dose.

Caffeine pharmacokinetic parameters for each cohort are shown in **Table 3**. We found caffeine Cmax, and Tmax were unaffected by methylliberine coadministration. However, methylliberine coadministration significantly increased t_1/2_ (14.7±5.8 vs 7.15±5.59 h), and AUC (70.8±36.9 vs 30.5±17.8 hxng/mL/mg). Moreover, methylliberine decreased caffeine oral clearance (CL/F, 17.1±7.8 vs 41.9±19.5 L/h) **Table3**.

**TABLE 3.** Summary of the statistical analysis of the pharmacokinetic parameters of methylliberine after single oral administration of a 100 mg dose of methylliberine alone and in co-administration with caffeine (150 mg)

| *Pharmacokinetic*<br>*Parameters of*<br>*Methylliberine* | *Geometric Mean* |  | *Geometric*<br>*mean ratio* | *90% CI* |
| --- | --- | --- | --- | --- |
|  | *Methylliberine*<br>*(100 mg) (Cohort II)* | *Methylliberine (100 mg) +*<br>*Caffeine (150 mg) (Cohort IV)* |  |  |
| C<sub>max</sub> (ng/mL) | 254 | 229 | 0.9 | 0.53-1.54 |
| T<sub>max</sub> (hours) | 1.0 | 1.0 | 1.0 | 0.84-1.85 |
| t<sub>1/2</sub> (hours) | 1.40 | 1.41 | 1.0 | 0.92-1.10 |
| AUC (h x ng/<br>mL/mg) | 6.69 | 8.26 | 1.2 | 1.04-1.47 |
| CL/F (L/h) | 149 | 121 | 0.81 | 0.68-0.97 |
| V<sub>d</sub>/F (L) | 303 | 246 | 0.81 | 0.69-0.96 |
| MRT (hours) | 2.31 | 2.39 | 1.0 | 0.93-1.15 |

After a single dose of caffeine 25 mg, the geometric mean t_1/2_ was 5.3 h, AUC was 26.78 ng·h/mL/mg, and CL/F was 37.34 L/h (**Table 4**). Compared with caffeine 100 mg, oral coadministration of methylliberine 100 mg with caffeine 150 mg resulted in an increase in the geometric mean t_1/2_ was 13.6 h, AUC was 63.98 ng·h/mL/mg, and in a decrease in the geometric mean CL/F was 15.63 L/h (**Table 4**). The geometric mean ratios for Cmax, Tmax, half-life, AUC, CL/F, Vd/F, and MRT on oral administration of caffeine (150 mg) versus caffeine 150 mg plus methlliberine 100 mg were 1.04, 1.63, 2.56, 2.39, 0.42, 1.07 and 2.55 respectively **Table 4**.

**TABLE 4.** Summary of the statistical analysis of the pharmacokinetic parameters of methylliberine after single oral administration of a 100 mg dose of methylliberine alone and in co-administration with theacrine (50 mg)

| *Pharmacokinetic*<br>*Parameters of*<br>*Methylliberine* | *Geometric Mean* |  | *Geometric*<br>*mean ratio* | *90% CI* |
| --- | --- | --- | --- | --- |
|  | *Methylliberine*<br>*(100 mg) (Cohort II)* | *Methylliberine (100 mg) +*<br>*Theacrine (50 mg) (Cohort V)* |  |  |
| C<sub>max</sub> (ng/mL) | 254.09 | 275.10 | 1.08 | 0.85-1.38 |
| T<sub>max</sub> (hours) | 0.81 | 0.83 | 1.03 | 0.74-1.42 |
| t<sub>1/2</sub> (hours) | 1.40 | 1.33 | 0.95 | 0.85-1.06 |
| AUC (h x ng/<br>mL/mg) | 6.69 | 6.35 | 0.95 | 0.82-1.10 |
| CL/F (L/h) | 149.40 | 157.50 | 1.05 | 0.91-1.22 |
| V<sub>d</sub>/F (L) | 302.60 | 302.30 | 0.99 | 0.90-1.11 |
| MRT (hours) | 2.31 | 2.39 | 1.03 | 0.93-1.15 |

Theacrine pharmacokinetic parameters were shown in **Table S3**. The study was not designed to determine the effect of caffeine and/or methylliberine co-administration on theacrine pharmacokinetics, viz., there was not an arm where subjects received only theacrine. However, based on our previous pharmacokinetic studies with theacrine and caffeine, it appears that methylliberine increased the half-life of theacrine by approximately two-fold **Table S3** ^9^.

## Discussion

In the United States, approximately 85% of people consume at least one caffeinated beverage per day with caffeine intake peaking between 50-64 years of age ^15^. The decline in caffeine consumption in adults 65 and older may reflect, in part, anecdotal reports of heightened susceptibility to caffeine-related adverse events with aging. Mostly, healthy adults with a moderate daily caffeine intake (≤400 mg) experience enhanced arousal, mood, and focus and experience little to no caffeine-related adverse effects^16,17^. Conversely, many individuals, particularly those with underlying medical conditions (*e*.*g*., hypertension), report an increased incidence of anxiety, nervousness, and jitteriness at caffeine doses >400 mg/day.^18^Variation in caffeine sensitivity has spurred the discovery of wider therapeutic index natural stimulant platforms that include a unique class of purine alkaloids known as methylurates^4,7,8,19^. For example, the methylurate theacrine exerts its pharmacologic effects via adenosine receptor modulation^20^, a property shared by its structural analog caffeine^21^. Intriguingly, however, the pharmacologic profile of theacrine appears distinct from caffeine in that it does not alter cardiovascular parameters (*e*.*g*., heart rate)^3,22,23^. For this reason, theacrine is frequently combined (“stacked”) with caffeine in energy, mood, and focus dietary supplements; unfortunately with little regard for pharmacokinetic and pharmacodynamic interaction potential.

To gain insight into the herb/drug interaction potential between methylxanthines and methylurates, we conducted a pharmacokinetic study in humans orally administered caffeine and/or theacrine^9^. When combined, caffeine diminished theacrine’s oral clearance (CL/F) without altering its half-life (t_1/2_ ∼Vd/CL), which suggested that the most likely mechanism for the observed interaction was that caffeine increased theacrine’s oral bioavailability (F). Similarly, in the current study, caffeine co-administration led to a modest, but significant, decrease in oral clearance (CL/F) and volume of distribution (Vd/F), as well as an increase in plasma area under the curve (AUC = (F*Dose)/CL) of methylliberine. However, methylliberine half-life, and by extension Vd and CL, were unaltered by caffeine. Again, the inference being that caffeine increased methylliberine’s oral bioavailability. These findings demonstrate that caffeine, acting through unknown molecular mechanisms, increases the bioavailability of methylurates such as theacrine and methylliberine. Because the vast majority of caffeine’s liver extraction ratio is attributable to cytochrome P450 1A2 (CYP1A2)^24^, it is tempting to speculate that methylurates are also CYP1A2 substrates. However, further research is needed to confirm this hypothesis.

In our previous study investigating the pharmacokinetic interaction potential between theacrine and caffeine, theacrine was found to have essentially no effect on caffeine bioavailability or clearance^9^. The inability of theacrine to increase caffeine bioavailability is not surprising as caffeine is a low extraction drug with an oral bioavailability approaching unity. However, the lack of an effect of theacrine on caffeine clearance is informative since it implied that theacrine, while it may be a CYP1A2 substrate, is not a clinically significant CYP1A2 inhibitor. In the current study, however, concomitant administration of caffeine and methylliberine led to significantly increased caffeine exposure (AUC), which was accompanied by commensurate decreases in half-life and oral clearance (CL/F). Interestingly, data from our previous study, although not designed to evaluate pharmacokinetic interaction potential, clearly showed that caffeine oral clearance (CL/F) was substantially lower than literature reports when co-administered as a cocktail also containing both theacrine and methylliberine^13^. The mechanism by which methylliberine reduced oral clearance (CL/F) of caffeine is unlikely related to increased bioavailability (F) considering the fact that caffeine’s bioavailability is complete and that caffeine’s oral volume of distribution (Vd/F) was unchanged.

A clue as to the potential mechanism by which methylliberine decreases the oral clearance (CL/F) of caffeine is provided by the fact that caffeine is a low hepatic extraction drug that is extensively metabolized (>90%) by CYP1A2 to the N3-demethylated metabolite paraxanthine ^25^. Hepatic clearance of low extraction drugs is approximated by multiplying the fraction of unbound drug (f_up_) and intrinsic clearance (CL_int_)^26^. Thus, a reduction in caffeine’s hepatic clearance is likely attributable to a reduction in intrinsic clearance, which reflects CYP1A2 activity. Thus, our data support the notion that methylliberine decreases the intrinsic clearance of caffeine through mechanisms likely including inhibition of CYP1A2. However, we cannot discount many other potential factors such as gender, race, genetic variation, disease, and exposure to inducers, which contribute to large interindividual variability in CYP1A2 activity and thus caffeine clearance ^25,27,28^. For example, caffeine’s plasma clearance is reduced in patients with liver cirrhosis, hepatitis B, and hepatitis C ^29,30^. Moreover, smoking stimulates caffeine clearance via CYP1A2 induction, whereas cessation of smoking decreases caffeine clearance ^28,31-33^. It is also puzzling that theacrine, which was administered at doses similar to methylliberine doses in this study, did not affect caffeine clearance in our previous study^9^.

In conclusion, methylliberine, a methylurate analog of caffeine, increased plasma exposure and half-life of caffeine following concomitant oral administration. The mechanism underlying this pharmacokinetic interaction is likely attributable to methylliberine inhibition of CYP1A2, which is a major determinant of intrinsic clearance, and thus hepatic clearance, of caffeine. Several important consequences, with regard to herb drug interaction potential, can be inferred from the data assuming reproducibility in larger more diverse populations. First, caffeine is commonly used as a probe drug to examine CYP1A2-mediated drug interactions^34^. Consequently, our data demonstrate that methylliberine has the potential to interact with other drugs whose elimination depends on CYP1A2. Secondly, methylurate pharmacology is still in its infancy, but early studies imply that methylxanthine and methylxanthine ligands differ in their affinity and selectivity for the adenosine A_1_ and A_2A_ receptors^20^, as well as, the sirtuin 3 receptor^35^. Ergo, additional pharmacology studies are needed to provide insight into the pharmacodynamic interaction potential between methylxanthines and methylurates.

## Data Availability

No additional data, outside that included in manuscript, was generated

## Acknowledgements

Funding for this work was provided by Compound Solutions, Inc.

## Tables

**Supplemental Table 1.** Caffeine pharmacokinetic parameters

| *Parameter* | *Cohort III* | *Cohort IV* |
| --- | --- | --- |
| $C_{\max}$ (ng/mL) | 439.8±139.5 | 458.1.3±93.5 |
| $T_{\max}$ (hours) | 1.43±0.99 | 1.9±1.3 |
| $t_{1/2}$ (hours) | 7.15±5.59 | 14.7±5.8 |
| AUC (h x ng/<br>mL/mg) | 30.5±17.8 | 70.8±36.9 |
| CL/F (L/h) | 41.9±19.5 | 17.1±7.8 |
| $V_d/F$ (L) | 350.6±147.9 | 315.5±76.4 |
| MRT (hours) | 10.1±7.9 | 21.6±9.6 |

**Supplemental Table 2.** Summary of the statistical analysis of the pharmacokinetic parameters of caffeine after single oral administration of a 150 mg dose of caffeine alone and in co-administration with methylliberine (100 mg)

| *Pharmacokinetic*<br>*Parameters of*<br>*Methylliberine* | *Geometric Mean* |  | *Geometric*<br>*mean ratio* | *90% CI* |
| --- | --- | --- | --- | --- |
|  | *Caffeine*<br>*(150 mg) (Cohort III)* | *Caffeine (150 mg) +*<br>*Methylliberine (100 mg) (Cohort IV)* |  |  |
| C<sub>max</sub> (ng/mL) | 428.14 | 445.00 | 1.04 | 0.93-1.16 |
| T<sub>max</sub> (hours) | 1.33 | 2.16 | 1.63 | 1.03-2.57 |
| t<sub>1/2</sub> (hours) | 5.30 | 13.60 | 2.56 | 2.06-3.19 |
| AUC (h x ng/<br>mL/mg) | 26.78 | 63.98 | 2.39 | 2.02-2.82 |
| CL/F (L/h) | 37.34 | 15.63 | 0.42 | 0.35-0.49 |
| V<sub>d</sub>/F (L) | 286.70 | 307.58 | 1.07 | 0.86-1.34 |
| MRT (hours) | 7.74 | 19.74 | 2.55 | 2.08-3.13 |

**Supplemental Table 3.** Theacrine pharmacokinetic parameters

| *Parameter* | *Cohort V* |
| --- | --- |
| C<sub>max</sub> (ng/mL) | 153.4±39.6 |
| T<sub>max</sub> (hours) | 1.9±1.4 |
| t<sub>1/2</sub> (hours) | 32.7±20.9 |
| AUC (h x ng/<br>mL/mg) | 130.9±71.6 |
| CL/F (L/h) | 9.5±4.1 |
| V<sub>d</sub>/F (L) | 358.1±101.8 |
| MRT (hours) | 48.1±29.5 |

